# Comparative Organ Disease Burden and Sequelae of Influenza and SARS-CoV-2 Infection: An Observational Study Using Real-World Data

**DOI:** 10.1101/2023.10.26.23297626

**Authors:** Istvan Bartha, Cyrus Maher, Victor Lavrenko, Keith Boundy, Elizabeth Kinter, Wendy Yeh, Davide Corti, Amalio Telenti

## Abstract

Infections with SARS-CoV-2 and influenza are associated with acute and post-acute complications and sequelae of many organ systems (i.e., disease burden). It is important to understand the global disease burden that associates with and follows acute infection in order to establish preventive and therapeutic strategies and to reduce the use of health resources and improve patient health outcomes. To address these questions, we utilized the National Covid Cohort Collaborative, which is an integrated and harmonized data repository of electronic health record data in the USA. From this database, we included in analysis 346,648 eligible SARS-CoV-2-infected patients, 78,086 eligible influenza infected patients, and 146,635 uninfected controls. We describe the disease burden that extends over 2-3 months following infection, and we quantify the reduction of disease burden by treatment. We identify a burden of disease following medically attended influenza that is comparable to that of medically attended SARS-CoV-2 infection. However, in contrast to SARS-CoV-2, influenza acute infection and disease burden are not responsive to antiviral treatment and thus remain as an unmet medical need. Focusing therapeutic strategies solely on the short-term management of acute infection may also underestimate the extended health benefits of antiviral treatment.

## Introduction

Infection with SARS-CoV-2 can result in a broad range of health consequences, from asymptomatic infection to respiratory insufficiency, hospitalization, and death. Additionally, severe cases of SARS-CoV-2 infection can lead to manifestations in various organs, herein referred to as “disease burden,” which can encompass pulmonary, cardiovascular, hematological, metabolic, gastrointestinal, kidney, mental health, musculoskeletal, and neurological disorders ^1^. These organ-related manifestations can present months after the acute infection and are also referred to as “sequelae” ^1,2^. In contrast, the disease burden and sequelae of influenza virus infection have received less attention (https://www.cdc.gov/flu/symptoms/symptoms.htm). Although the pulmonary complications of influenza are well-understood, influenza virus infection can involve numerous organ systems, as described in an extensive review by Sellers et al.^3^ in 2017. That review highlighted that the recognition of these extra-pulmonary complications is crucial in determining the actual health consequences of influenza infection. In particular, severe forms of influenza infection such as H7N9 avian flu, can result in long-term disability and psychological impairment that persist even two years after discharge from the hospital ^4^. Empirical estimates of the full global influenza disease burden are complicated by challenges of associating non-respiratory outcomes with influenza, especially when laboratory confirmation is rare ^5^. Therefore, quantifying the impact of disease beyond the acute infection episode is critical for establishing the full burden of SARS-CoV-2 and influenza infections on patients and on the health system.

Effective treatments for SARS-CoV-2 are now available, and randomized clinical trials have demonstrated their ability to reduce hospitalization, severe disease, and death rates. Recent publications have shed light on the SARS-CoV-2 disease burden and time profile after acute infection ^1,2^ and the impact of Paxlovid treatment on post-acute sequelae of COVID-19 ^6^. Vaccination for infuenza is also associated with measurable diminution of the global organ disease burden ^5^. However, there has been considerable debate regarding the benefits of antiviral treatment against acute influenza infection ^7^. Here we compare these two viral infections in terms of burden of acute and post-acute disease burden and in their response to current antiviral therapeutics.

## Materials and Methods

### Definition of the study group

The National Covid Cohort Collaborative (https://ncats.nih.gov/n3c) is an integrated and harmonized data repository of electronic health record data. The database includes data of 7 million SARS-CoV-2-infected individuals and 10 million controls, including individuals that had influenza. We selected individuals who satisfied predetermined inclusion and exclusion criteria to allow assessment of the efficacy of antiviral treatment. Inclusion criteria were: availability of age and body mass index [BMI], because their importance in disease severity and requirement for balancing of the risk. Exclusion criteria were: immunosuppression, reinfection within 6 months, hospitalization within 7 days of documented infection, death within 7 days documented infection and treatment delayed by more than 5 days after infection. These exclusion criteria were used to allow for optimal assessment of the efficacy of treatment. In this paper, we use the term medically attended infections to emphasize that we only observe infections that result in contact with the health system.

We assessed the use of paxlovid and monoclonal antibodies in the treatment of SARS-CoV-2, as well as the use of oseltamivir and baloxavir for the treatment of influenza. To serve as controls in the estimation of treatment efficacy we selected untreated Covid infected individuals from the eligible set and used all eligible influenza infected individuals for the analysis. For the assessment of the difference in the incidence of diseases between infected and uninfected people we matched uninfected individuals from the eligible set.

### Elaboration of concepts

We used Supplementary Table 13 from the work of Xie et al ^2^ to establish 18 coarse grained organ specific disease categories. Each category has 1-5 concrete disease manifestations. We mapped these concrete, individual diseases to Observational Medical Outcomes Partnership (OMOP, https://ohdsi.org/omop/) concepts both manually and automatically. The automatic mapping was done by tokenization and full text search, the manual mapping was done by utilizing the ‘Concept Set Browser’ in N3C Enclave (https://covid.cd2h.org/). We defined total disease burden as the sum of all disease categories except the muscoskeletal and psychological disease categories.

### Matching

We analyzed one-to-one matched pairs of individuals to minimize biases in the estimates. We used maximal bipartite graph matching on the bipartite max-radius nearest neighbor graph of treated and untreated individuals. In that graph an edge connects a treated and an untreated individual if the distance between them is less than a threshold, which is a hyperparameter set to 0.2. We choose this value to be as small as possible while still matching more than 85% of the smaller partition of the graph. We use L2-norm as distance function on a space of the following variables: logit of propensity score (z-transformed), age (z-transformed), BMI (z-transformed), sex (binary).

Propensity scores were estimated with gradient boosted trees on multiple variables ^8^. The trees were built on 237 variables including demographics, pre-Covid burden of organ-specific disease categories, and manually extracted laboratory measurements, conditions, or observations.

### Statistics

We estimated the odds ratio from the matched contingency tables either directly or by multivariate conditional logistic regression ^9^. We used the same variables in this regression as were used to compute the Euclidean distances for matching. When estimating odds ratios in the first quarter after infection we defined as “novel diagnosis” any disease reports that were not recorded during the 24 months before the infection. We report the confidence intervals from the conditional logistic regression if that converged, otherwise we report the confidence intervals computed directly from the matched contingency table. To quantify the excess disease burden upon infection, we report the odds ratio of having a novel diagnosis of a disease during the acute and post-acute period in infected people and a chosen control period of the same length and same calendar era of uninfected people. To quantify efficacy, we report the odds ratio of having a novel diagnosis of a disease in the post-acute period between the treated and untreated populations.

## Results

We identified 7 million SARS-CoV-2 positive and 400 thousand influenza positive individuals in the N3C dataset. A small subset of those were found eligible based on the pre-defined exclusion and inclusion criteria. In particular we could not establish the age (0.45 million missing) and the BMI (4 million missing) of numerous SARS-CoV-2 positive individuals. In addition, about 1.7 million had no laboratory evidence of infection, or were hospitalized shortly after infection (0.3 million), thus limiting the possibility to assess drug response.

We aimed at comparing disease management across the two viral infections over a contemporary period and with a general availability of antiviral treatment. Thus, we focused our analysis on the periods of the delta and omicron SARS-CoV-2 variants of concerns, that is beginning from 2021 May until 2022 November. In the case of influenza, we focused the analysis on the year 2022. During these periods, we included as final analysis sets 346,648 patients infected with SARS-CoV-2 (including 68,615 eligible treated patients), 78,086 patients infected with influenza (including 26,436 eligible treated patients), and 146,635 matched uninfected controls. **Suppl. Table S1** depicts the number of patients treated with various treatment options and further stratifies the numbers by the calendar period in which the infection occurred.

### Disease burden

The N3C dataset is normalized into the OMOP electronic health record schema. We used the 18 coarse grained organ specific disease categories (cardiac disorders, central nervous system, depression, dermatitis, diabetes, dyspnea, gastrointestinal, hypertensive renal disease, hypoxia, kidney, neoplasms, pulmonary disorders, thrombotic disorders and ventilation support) described in Methods, **Suppl. Table S2**. The final list of OMOP concepts, coarse grained disease categories and the frequency of the given concept in the analysis cohort for a final total of 805,477 treated or untreated SARS-CoV-2 or influenza infected individuals) is presented in **Suppl. Table S3**.

### Bias in treatment allocation

We computed descriptive statistics of age, BMI, sex, and two-year pre-infection burdens of 18 organ-specific disease categories (**Table 1**). We found that individuals receiving treatment have different baseline demographics and disease burden which reflect treatment allocation bias. This also applies uniformly to individual disease categories: baseline rates of anemia, diabetes, dysrhythmias, gastrointestinal problems, ischemic heart disease, kidney disease, thrombotic disorders were twice more frequent in the treated population.

**Table 1.** Baseline characteristics of study participants. Descriptive statistics of age, sex, BMI, and burden of diseases stratified by treatment. Values established for the 1-24 month period preceding acute infection.

|  | Treated SARS-CoV-2<br>(n=68,615) | Treated Influenza<br>(n=26,436) | Uninfected<br>(n=146,635) | Untreated SARS-CoV-2<br>(n=278,033) | Untreated Influenza<br>(51,650) |
| --- | --- | --- | --- | --- | --- |
| **Demographics [mean (sd)]** |  |  |  |  |  |
| Age | 56.26 (17.76) | 29.25 (24.59) | 44.17 (24.11) | 38.70 (22.82) | 23.25 (21.74) |
| BMI | 31.29 (7.96) | 25.19 (8.66) | 27.37 (8.26) | 27.39 (8.77) | 23.84 (8.52) |
| Female | 0.58 (0.49) | 0.56 (0.50) | 0.57 (0.49) | 0.58 (0.49) | 0.55 (0.50) |
| **Pre-existing burden [prevalence 1-24 months before infection, %]** |  |  |  |  |  |
| Cardiac disorders | 1.47% | 0.85% | 1.16% | 0.69% | 0.35% |
| Central nervous system | 1.42% | 0.80% | 1.21% | 0.75% | 0.42% |
| Depression | 1.82% | 1.00% | 1.63% | 1.21% | 0.79% |
| Dermatitis | 1.00% | 1.47% | 0.86% | 0.97% | 1.44% |
| Diabetes | 4.98% | 2.47% | 3.15% | 2.58% | 1.40% |
| Dyspnea | 2.98% | 2.01% | 2.20% | 1.60% | 1.27% |
| Dysrhythmias | 2.57% | 1.69% | 2.09% | 1.33% | 1.00% |
| Gastrointestinal | 4.46% | 2.71% | 3.46% | 2.55% | 1.97% |
| Hypoxia | 0.36% | 0.59% | 0.41% | 0.22% | 0.24% |
| Ischemic heart disease | 2.08% | 0.99% | 1.53% | 0.90% | 0.50% |
| Kidney | 1.06% | 0.73% | 1.02% | 0.62% | 0.29% |
| Neoplasm | 3.67% | 1.34% | 2.62% | 1.63% | 0.84% |
| Pulmonary | 0.53% | 0.36% | 0.44% | 0.25% | 0.15% |
| Thrombotic disorders | 0.37% | 0.25% | 0.33% | 0.19% | 0.10% |
| Ventilation support | 0.28% | 0.34% | 0.40% | 0.20% | 0.15% |
| Any of above | 9.72% | 5.80% | 7.57% | 5.69% | 3.98% |

### Disease burden dynamics before and after medically attended SARS-CoV-2 and influenza infection

In a first step, we computed disease burden across disease categories before and after medical attended SARS-CoV-2 and influenza infections in weekly strata. We defined weekly disease burden as the fraction of individuals having at least one disease record in a given period divided by the total number of individuals. We computed these time-resolved burdens in all treated and untreated SARS-CoV-2 and influenza cases, **Suppl. Figure 1.** In general, we found that the apparent disease burden peaks in the acute phase of infection and returns to pre-covid baseline levels by the second quarter.

We wanted to quantify the excess burden of disease after medically attended SARS-CoV-2 or influenza infection, which we do by comparing the events happening in the months after the infection to events happening in matched uninfected patients during a control period. Comparing the post-acute quarter of infected patients to an arbitrary three months of matched uninfected patients shows a significant increase in burden of almost all analyzed disease categories, with the sole exception of neoplasms (**Figure 1A and B, Suppl. Table S4**). However, comparing post-acute events to a period after an arbitrary non-infectious medical attended event shows that while the increase in the rates of mortality, hypoxia, dyspnea and thrombosis in the first three months after infection remain, there is a decrease in cardiac disorders, diseases of the central nervous system, diseases of the gastrointestinal tract and neoplasms (**Figure 1C and D, Suppl. Table S4**). This indicates that a medically attended event – infectious or non-infectious – may associate with an increase in disease burden.

**Figure 1.**
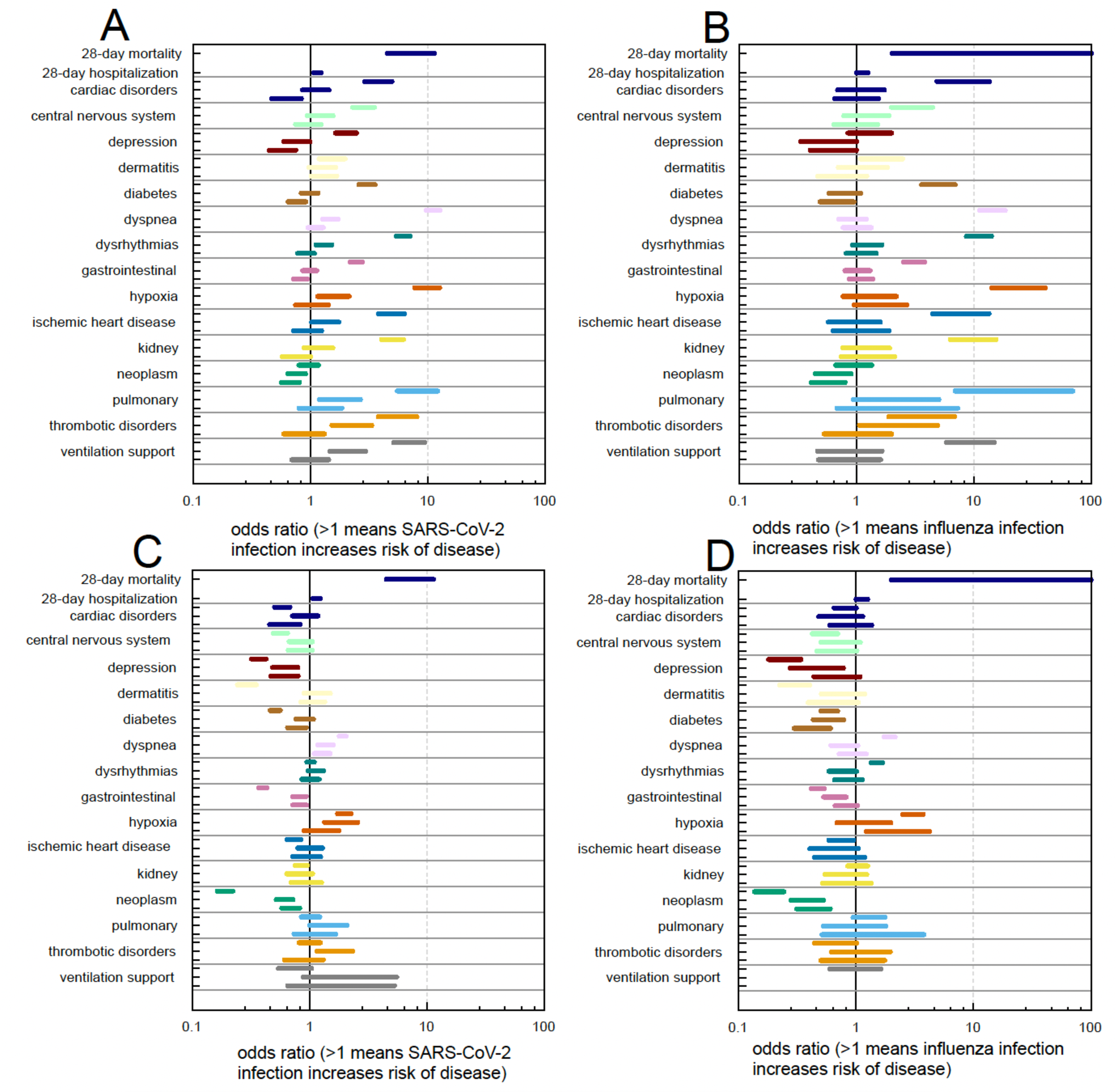
Excess burden in the post-acute period. Estimated odds ratios for having a new disease manifestation during acute infection and up to 3 months after infection compared to uninfected patients during a selected control period. **Panel A** (SARS-CoV-2) and **B** (influenza), control period is an arbitrary period not necessarily linked to a medical contact. **Panel C** (SARS-CoV-2) and **D** (influenza), control period is a period marked by a medical contact of a different nature (non-viral). For each organ manifestation or disease, the figure provides three estimates that correspond to three consecutive periods/months: days 0-28, 29-56 and 57-84.

In summary, the measurement of disease burden is highly dependent on the choice of control/reference group and period. It is thus critical to differentiate a “re-call” phenomenon, when a list of active and historical diagnoses is created during a medically attended event, from a true association of an acute infection with new multiorgan disease diagnoses. The response to antiviral treatment – as described in the sections below – is determinant to establish the nature of the newly described disease burden.

### Effects of treatment on mechanical ventilation and death

We estimated the effect of treatment against 28-day all-cause mortality and 28-day ventilation support upon infection. We controlled for confounding by using 1 to 1 matching of untreated individuals to treated individuals. We based this matching on propensity scores, age, sex, and BMI. We find that our estimates agree with previously published effects from SARS-CoV-2 clinical trials. The odds ratio of anti-SARS-CoV-2 treatment against mortality in this calendar period is 0.15-0.32 (equivalently 68-85% protection), **Figure 2A, Suppl. Table S4**. The odds ratio of anti-influenza treatment against mortality is not significant (**Figure 2B, Suppl. Table S4**).

**Figure 2.**
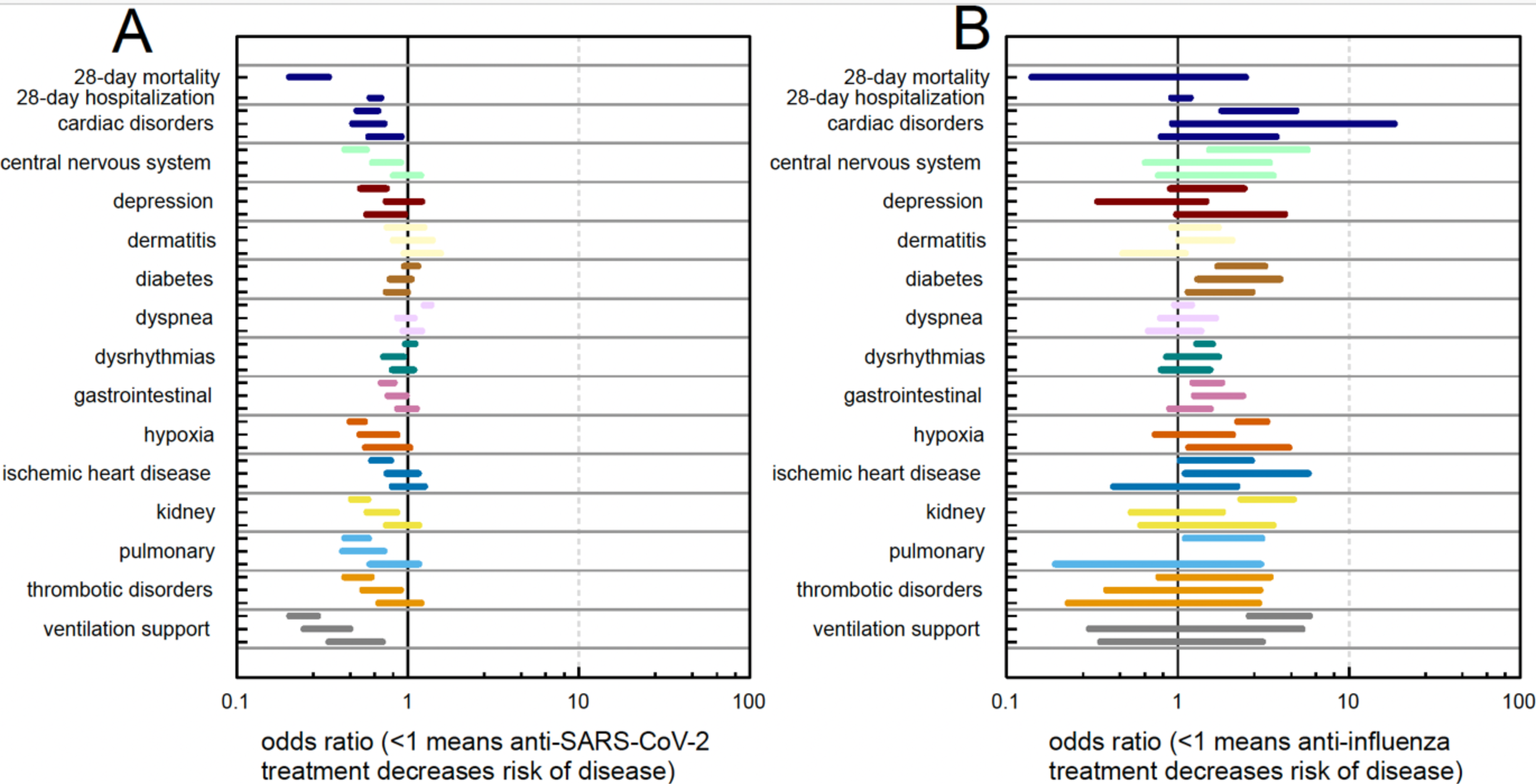
Effectiveness of antiviral treatment. Estimates, odds ratios, for the effect of anti-SARS-CoV-2 (**panel A**) or anti-influenza (**panel B**) treatment against severe manifestations of acute infection and acute and post-acute disease burden. For each organ manifestation or disease, the figure provides three estimates that correspond to three consecutive periods/months: days 0-28, 29-56 and 57-84.

### Effects of treatment on disease burden

We estimated odds ratios on disease burden between treated and untreated groups. We measured a significant protective effect of anti-SARS-CoV-2 treatment against disease of the central nervous system, cardiac disorders, pulmonary disease, kidney disease and against thrombotic disorders. Effects of anti-SARS-CoV-2 treatments against diabetes were inconclusive while the estimated odds ratio of dyspnea in the first acute month is significantly higher than one (**Figure 2A, Suppl. Table S5)**, likely the result of treatment allocation bias towards to the sickest patients. In contrast, we failed to establish a protective effect for anti-influenza treatment across organ disease categories (**Figure 2B, Suppl. Table S5)**. Actually, we observed increased disease burden among individuals treated with anti-influenza agents. This is a strong indication of treatment allocation bias towards the sickest patients.

In summary, treatment of SARS-CoV-2 reduces severity of the acute event and limits non-respiratory disease burden. In contrast, treatment of influenza appears ineffective possibly because of late prescription, treatment allocation bias to the sickest, or inherent limitation of the drugs.

## Discussion

Reporting on treatment of SARS-CoV-2 and influenza infection has generally focused on the protection from hospitalization, severe disease and death. However, there is full awareness of the multiorgan consequences of SARS-CoV-2 infection. Here we show that there is an increase in disease burden across multiple organ systems that only returns to baseline 3 months post-acute infection. Less reported, we show that infection with influenza virus results in a comparable pattern of acute and post-acute organ disease burden – including the occurrence of thrombotic events that are thought of as paradigmatic of COVID-19 but that we now observe in influenza infection. We also show that treatment with direct acting antiviral agents and biologicals diminishes disease burden up to 3 months following acute SARS-CoV-2 infection. In contrast, we cannot establish a protective effect of treatment of influenza infection against excess disease burden in the real-world usage of oseltamivir.

The magnitude of the SARS-CoV-2 pandemic facilitated the observation and recording of the large number complications of infection. What was notable from studying a similarly sized population collected as controls in the context of the NCATs was the possibility to analyze approximately 78,000 influenza infection events. The global rates and pattern of organ disease burden was similar for the two viruses. Where the two viruses differed was on the observed effectiveness of treatment. Treatment of SARS-CoV-2 resulted in substantial benefit on acute (hospitalization, severity and death) and on acute and post-acute organ disease burden. In contrast, treatment of influenza – mainly oseltamivir – was not associated with measurable efficacy. This is consistent with the Cochrane review of the limited efficacy of neuraminidase inhibitors ^7^ for the treatment of influenza. However, it should be underscored that the real-world data in the present study indicated that disease burden increased ahead of influenza diagnosis – suggesting delayed testing from time of symptom onset and probably late treatment of influenza. Influenza testing is not available at-home, is done by health care providers and time to receive results can be longer. This stands in contrast to the wide availability of and timely results of SARS-CoV-2 testing during the pandemic.

As indicated above, real world data analytics is fraught with confounding and biases. True for both viral infections, people who received antiviral treatment were significantly older and with more morbidity than non-treated individuals, which reflects considerable treatment allocation bias ^10^. The main difficulty is achieving optimal matching of individuals that did or did not receive treatment because they were profoundly different across demographics and underlying organ morbidity prior to acute viral infection.

In our study we do not observe the totality of SARS-CoV-2 and the totality of influenza infections. In fact, the probability of diagnosing a SARS-CoV-2 infection differs substantially from the probability of diagnosing influenza, and they both differentially depend on the severity of presented symptoms. This yields a bias in the observed, medically attended cases of infections, which leads to some limits to the interpretations of our comparative results. All our results are conditioned on medical contact.

Despite the attention to post-acute sequelae of SARS-CoV-2 infection (PASC, long-covid syndrome), in the broadest terms defined as persistence of a broad set of disease manifestations 3 months after acute infection (https://www.who.int/europe/news-room/fact-sheets/item/post-covid-19-condition, and https://www.covid.gov/longcovid/definitions), we observed a global return to pre-acute infection organ disease burden by the end of the first 3 months. Thus, the study design is not specific or sensitive to the identification of patients that could be classified as having PASC. Nor could we assess the role of treatment for this late complication of SARS-CoV-2 infection.

We find that excess disease burden is not definable on its own and the baseline to which the excess is measured is an important variable. Measured post-acute burden is higher if compared to a disease-free baseline period, than if compared to a period which starts with a medical contact of any nature. This could have been interpreted as the excess in disease burden after viral infection reflecting the recall of previous and pre-existing diseases rather than a true consequence of acute viral infection. The effect of treatment is a powerful means of answering to this question: reduction of various non-pulmonary diseases by anti-SARS-CoV-2 provide evidence that such the burden of disease is truly associated with acute infection rather than a simple recall of preexisting disease.

In summary, the study calls for increasing attention to the value of treatment for the prevention of complications and disease burden of SARS-CoV-2 infection. It also indicates that there is a need for more effective treatments for influenza or prophylactic therapies to prevent infection, as well as earlier influenza diagnostics and more agile prescription – hopefully leading to treatment responses more in line with those obtained for SARS-CoV-2. In addition to the treatment considerations, further research should focus on understanding the health resource utilization and health economic burden associated with complications of influenza given that the excess organ disease burden translates in increasing use of health resources and visits. This would enable identification of potential cost-savings and improved patient outcomes from both earlier treatment and prevention infection, especially in high-risk individuals.

## Data Availability

Access to data through the NCATS N3C Data Enclave covid.cd2h.org/enclave

https://ncats.nih.gov/n3c

## Supplementary materials

**Supplementary Figure 1.**
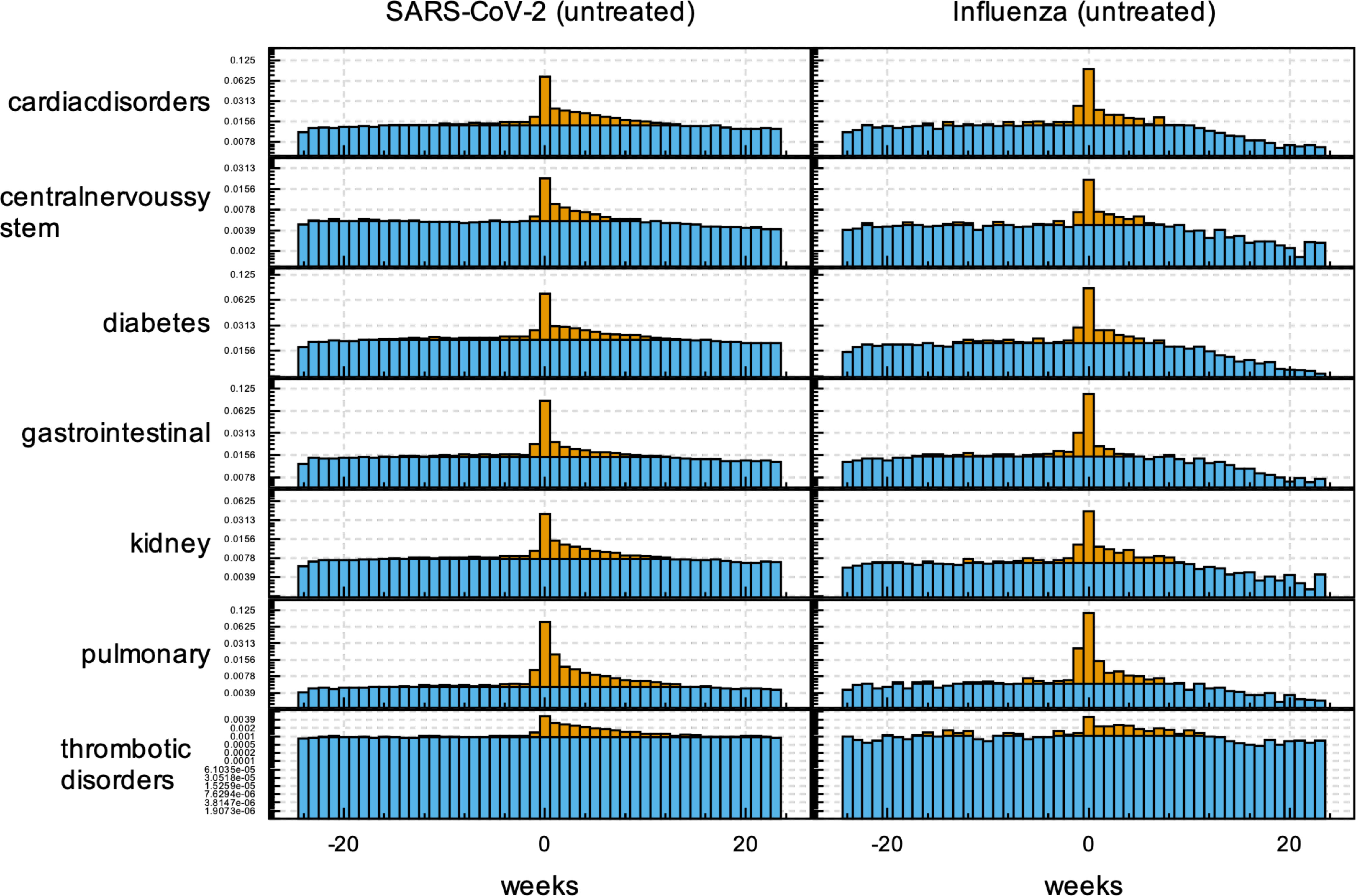
Comparative disease burden for SARS-CoV-2 and influenza infection. Weekly rates of occurrences of disease categories centered around SARS-CoV-2 and influenza infection (blue). Burden over baseline is depicted in yellow.

**Supplementary Table S1.** Treatment of viral infection. Count of SARS-CoV-2 or influenza infected individuals in the analysis group who were treated against Covid or influenza infection, or received no treatment in four non overlapping time periods (before Delta, Delta, Omicron BA1/BA2, Omicron BA5).

|  | All time | 2020-01-01<br>-<br>2021-05-01 | 2021-05-01 -<br>2022-01-01 | 2022-01-01 -<br>2022-11-01 | 2022-11-01 -<br>2023-01-01 |
| --- | --- | --- | --- | --- | --- |
| **paxlovid** | 37'989 | <20 | 1'568 | 24'880 | 11'524 |
| **sotrovimab** | 4'684 | <20 | 2'334 | 2'345 | <20 |
| **bebtelovimab** | 9'075 | <20 | 531 | 6'968 | 1'576 |
| **bamlanivimab** | 11'743 | 6'418 | 4'025 | 1'300 | <20 |
| **casirivimab** | 29'308 | 4'643 | 17'992 | 6'672 | <20 |
| **baloxavir** | 752 | 410 | 34 | 160 | 148 |
| **oseltamivir** | 107'370 | 60'952 | 3'879 | 22'363 | 20'176 |
| **peramivir** | <20 | <20 | <20 | <20 | <20 |
| **zanamivir** | <20 | <20 | <20 | <20 | <20 |

**Supplementary Table S2.** Disease keywords. Organ specific coarse disease categories and finer grained individual diseases used to establish disease burden.

| Category | Keywords |
| --- | --- |
| Cardiac disorders | Cardiac arrest |
| Cardiac disorders | Cariogenic shock |
| Cardiac disorders | Heart failure |
| Cardiac disorders | Myocarditis |
| Cardiac disorders | Non-ischemic cardiomyopathy |
| Cardiac disorders | Pericarditis |
| Central Nervous System | Alzheimer's disease |
| Central Nervous System | Central Venous thrombosis |
| Central Nervous System | Memory problems |
| Central Nervous System | Stroke |
| Central Nervous System | Transient cerebral ischemic attack (TIA) |
| Central Nervous System | Transverse myelitis |
| Depression | Depression |
| Dermatitis | Dermatitis |
| Diabetes | Antihyperglycemic |
| Diabetes | Diabetes |
| Diabetes | HbA1c >6.4 |
| Dyspnea | Dyspnea |
| Dysrhythmias | Atrial Flutter |
| Dysrhythmias | Atrial fibrillation |
| Dysrhythmias | Bradycardia |
| Dysrhythmias | Tachycardia |
| Dysrhythmias | Ventricular arrhythmias |
| Gastrointestinal | Acute gastritis |
| Gastrointestinal | Acute pancreatitis |
| Gastrointestinal | Appendicitis |
| Gastrointestinal | Cholangitis |
| Gastrointestinal | Cholecystitis |
| Gastrointestinal | Cholelithiasis |
| Gastrointestinal | Exocrine pancreatic insufficiency |
| Gastrointestinal | Functional dyspepsia |
| Gastrointestinal | Gastroesophageal reflux disease (GERD) |
| Gastrointestinal | Gastroparesis |
| Gastrointestinal | Hematemesis |
| Gastrointestinal | Malabsorption |
| Gastrointestinal | Melena |
| Gastrointestinal | Neurogenic bowel |
| Gastrointestinal | Paralytic ileus |
| Gastrointestinal | Peptic ulcer disease |
| Gastrointestinal | Ulcerative colitis |
| Hypoxia | Hypoxemia |
| Hypoxia | Hypoxia |
| Ischemic heart disease | Acute coronary disease |
| Ischemic heart disease | Angina |
| Ischemic heart disease | Ischemic cardiomyopathy |
| Ischemic heart disease | Myocardial infarction |
| Kidney | Acute kidney injury |
| Kidney | Chronic Kidney Disease |
| Neoplasm | Neoplasm |
| Pulmonary | Interstitial lung disease |
| Pulmonary | emphysema |
| Thrombotic disorders | Deep vein thrombosis |
| Thrombotic disorders | Pulmonary embolism |
| Thrombotic disorders | venous thrombotic embolism |
| Ventilation support | Ventilation support |

**Supplementary Table S3. Disease concepts mapped to the database schema and patient population.** Organ specific coarse disease categories, as well as selected signs and symptoms, all mapped OMOP concepts and the number of individuals with at least one such OMOP concept record in the analysis group.

**Supplementary Table S4. Estimated odds ratio of treatment types on subcategories of disease concepts.**

